# Connecting in COVID 19: Neurology tele-follow-up experience

**DOI:** 10.1101/2020.07.13.20153171

**Authors:** Deepti Vibha, MV Padma Srivastava, Kameshwar Prasad, Manjari Tripathi, Achal Kumar Srivastava, Rohit Bhatia, Mamta Bhushan Singh, VY Vishnu, Roopa Rajan, Rajesh Kumar Singh, Anu Gupta, Animesh Das, A Elavarasi, MR Divya, Bhargavi Ramanujam, A Shariff

## Abstract

**Introduction:** The lockdown due to COVID-19 pandemic led to temporary closure of routine hospital services. This prompted the initiation of teleconsult follow-up in our department. The study outlines the experience of tele-follow-up at a tertiary care teaching hospital in India, and the perspective of neurologists about this novel approach.

**Methods:** The tele-follow-up was started from 26th March 2020. Data of follow up appointments was provided by the medical record section. The faculty and senior residents conducted the tele-follow-up. Communication was made via voice calls. The data for initial ten days was analyzed to find the utility and experience of the new service.

**Results:** In the initial ten working days, data of 968 patients was provided for tele-follow-up. A successful communication was made in 50.3% patients (contact with patients: 27.7% and family members 22.6%). The phone numbers which were not contactable/invalid/not available constituted 36.8% of the data. A total of 35 faculty and residents conducted the tele-follow-up. The utility of tele-follow-up was perceived as good by 71.4% of neurologists. Majority of neurologists (71.4%) observed that >90% of patients were continuing medications. Patients outside the city constituted 50-75% of the list. The survey revealed that all neurologists felt the need to continue tele-follow-up for far off stable patients post lock down and resumption of regular outpatient services.

**Conclusion:** The survey established the feasibility and utility of teleconsult for follow up of patients with neurological diseases who were attending the regular outpatient services before the lock down.

## Introduction

While telemedicine is being used in India^1^, it has mainly focused on educational needs and reaching out to remote areas. The telemedicine services at our institute are largely providing educational services.^2^ Teleconsultation and tele-follow up have been out of the realm of tertiary health care professionals and systems are overloaded with conventional outpatient services with footfalls of several hundred per day.

While the lockdown period was a challenge for patients with neurological diseases, it provided an opportunity for us to reach out to them. Given the nature of our institution as a tertiary care referral center, patients often visit us from faraway places across the length and breadth of India. With over 1200 million telephone subscribers in India and a tele density of 91.8%, teleconsultation was considered as an option to provide continuity of care^3^. The initial feasibility and administrative issues of doing tele-follow up were settled by discussion with the authorities. The aim was to examine the feasibility and applicability of tele-follow-up of Neurology patients.

## Materials and Methods

The tele-follow-up services were started in the department of Neurology, All India Institute of Medical Sciences (AIIMS), New Delhi, India after a consensus by faculty members and with approval of the administration. The telemedicine guidelines by the Ministry of Health and Family Welfare^3^ was the reference document for the conduct of the teleconsultation services. The Neurology outpatient department (OPD) services has an online appointment system for new as well as follow-up patients. In addition, once the patient is on follow up, there is a hardcopy file for each patient which is maintained by the record section. For the conduct of tele-follow-up, the record section provided the details of the follow up patients along with their files, a day prior to the OPD day. Dedicated smartphones and hospital numbers were provided by the Institute authorities for conducting the tele-follow-up. The routine OPD timings were utilized to call up the patients who were already under our follow-up, as the initial run.

Each faculty/resident would call the patient and introduce himself/herself and confirm the number. After explaining the objectives of the call and obtaining a telephonic consent, the verbal consent was recorded in the patients file. The call would proceed based on a flexible approach about the details provided by the patient. The information about consent, the details about the progress and prescription was noted in the OPD file. In addition, the time spent during each consultation was noted. The occasions where the patient contact numbers being wrong; not answered after repeated attempts or no number listed were noted. Ethical clearance was taken from the Institute Ethics committee (IEC-301/17.04.2020).

Since the objective of initiating tele-follow-up services was to find out its utility, we also asked the faculty/residents conducting the teleconsultation services about their experience through a questionnaire.

## Results

The data was collected from 26^th^ March 2020 to 9^th^ April 2020 (10 working days). A total of 968 patients were seen in 10 days (average of 96.8 patients per day). We could contact the patients (27.7%) or their families (22.6%) about half the times. An interesting observation amongst the group where we could contact the family members was that some of them were local relatives or relatives living in bigger cities while the patient was living distantly in a smaller town/village. Numbers where no one answered the call (24.3%) and incomplete information by faculty/resident (12.9%) constituted the most common causes of inability to acquire information. Non-availability of either the patient’s phone numbers (2.4%) or OPD files (3.7%) constituted other reasons for our inability to contact patients. However, some patients were contacted without OPD files as well (table 1). The patients’ diagnosis, results of prior investigations and records of previous follow-up were obtained from the patients’ files prior to making the call.

**Table 1:** Proportion of patients who could be contacted by tele-consult.

|  | Number (n=968) (n(%)) |
| --- | --- |
| Contacted | 487 (50.3) |
| Communication with the patient | 268 (27.7) |
| Communication with a family member or relative | 219 (22.6) |
| Not contacted | 481 (49.7) |
| No one picked up on the number provided | 235 (24.3) |
| Incomplete data provided by faculty/trainees | 125 (12.9) |
| Invalid number, could not be contacted | 60 (6.2) |
| No hardcopy file available | 34 (3.5) |
| No number available in the database | 27 (2.8) |

The study adopted the simplest method of telephonic conversation in the group of patients who had already been on follow up in the department. These patients had fairly clear diagnosis and the OPD files documented the last visit details. Confirmation of ongoing treatment and minor changes based on the feedback of patients was the most common intervention. Advice to seek in-person visit to a local practitioner in case of doubt was advised when indicated.

The senior residents who were posted in COVID 19 wards, intensive care units (ICUs) and rotatory off did not participate in tele-follow-up. However, some faculty members could come on the scheduled day and time. Thus, 13 of the 15 faculty members, 21 of the 37 senior residents, and one stroke fellow provided tele-follow-up services at different times. The median time for conversation was two minutes (IQR: 1.19;3.07).

After ten days, we sent a questionnaire to all 35 Neurology faculty and trainees about their experience (table 2). Most opined that the tele-follow-up was useful (good:71.4%, fair:14.3%), and they were able to fulfill the purpose of conversation (good:57.1%, fair: 37.1%). Although there was some difficulty in getting the refill medication due to lockdown (sometimes: 34.3%, frequently:31.4%), most of the patients were continuing medications (>90% of the time: 71.4%, 71-90% of the time: 17.1%). Medical expenses in India is usually borne out of pocket, and some poor patients who depended on free hospital dispensary did express their constrains, although the exact numbers were not collected in this study. When the perceived level of satisfaction of the patients was asked, majority of the faculty and residents (85.7%) felt that their patients would be fairly satisfied. While the primary purpose of the tele-follow-up was to provide medical advice, a majority were involved in reassuring and allaying anxiety of the patients as well (77.1%). 62.9% of the respondents had 50-75% of their patients outside Delhi. Apart from the primary medical problem, replying to the concerns about pandemic, mental health, the prospects of resumption of OPD services were answered by 60.0% of Neurologists. Additional smartphone applications like images/videos of patients were not used by majority (74.3%). All the respondents felt that the tele-follow-up is feasible and useful for patients who are stable and stay far and should be continued beyond the lockdown period as well.

**Table 2:**
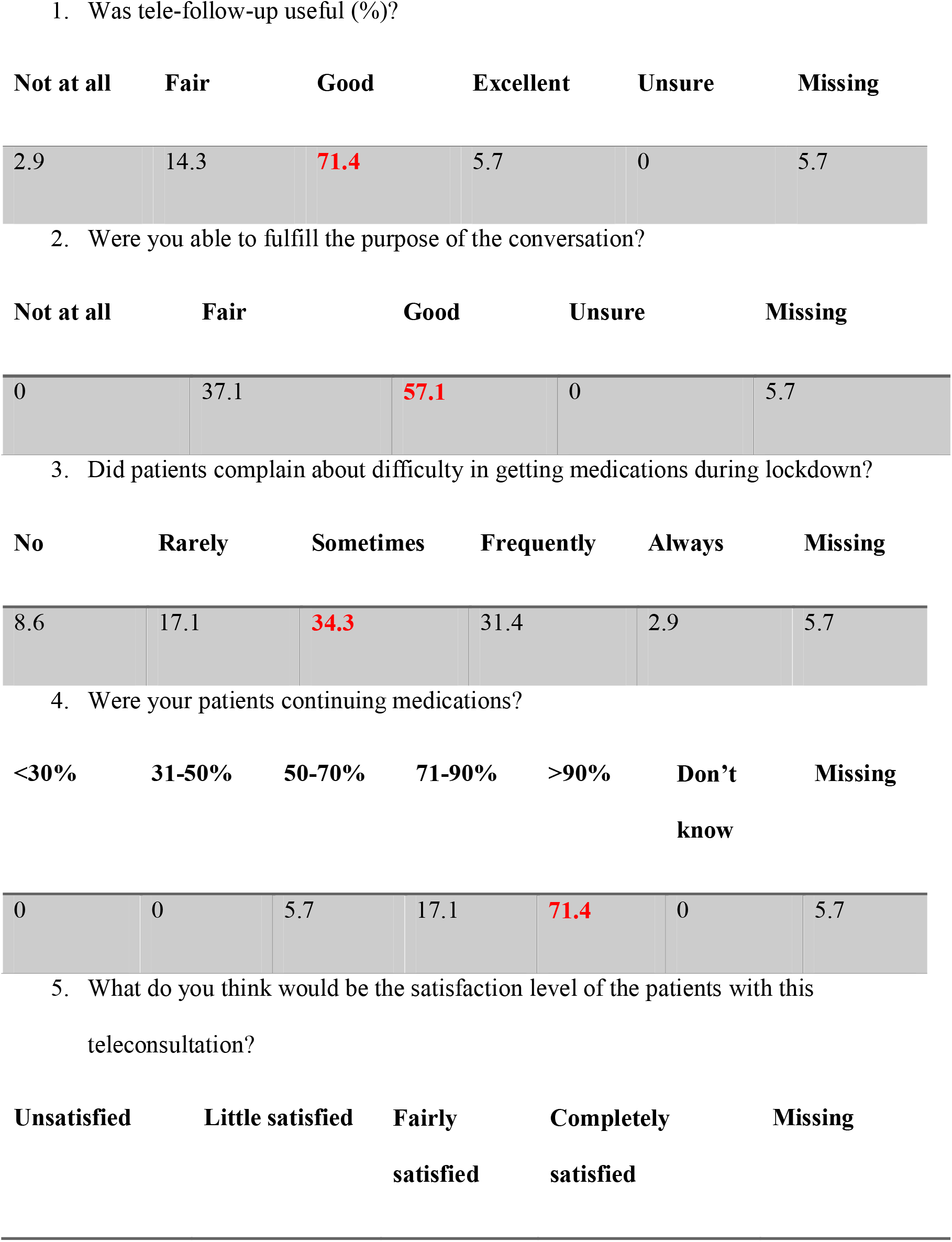
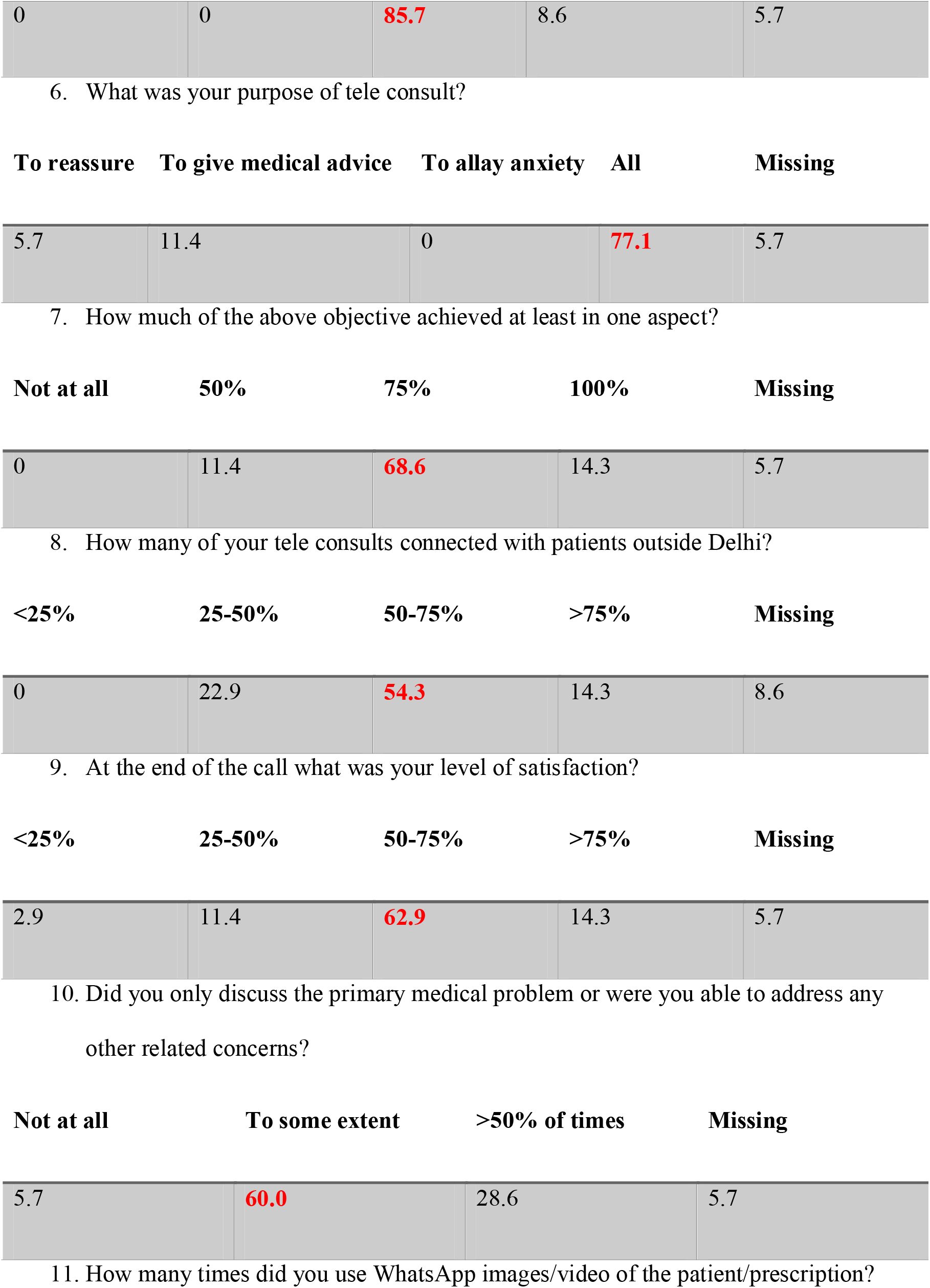
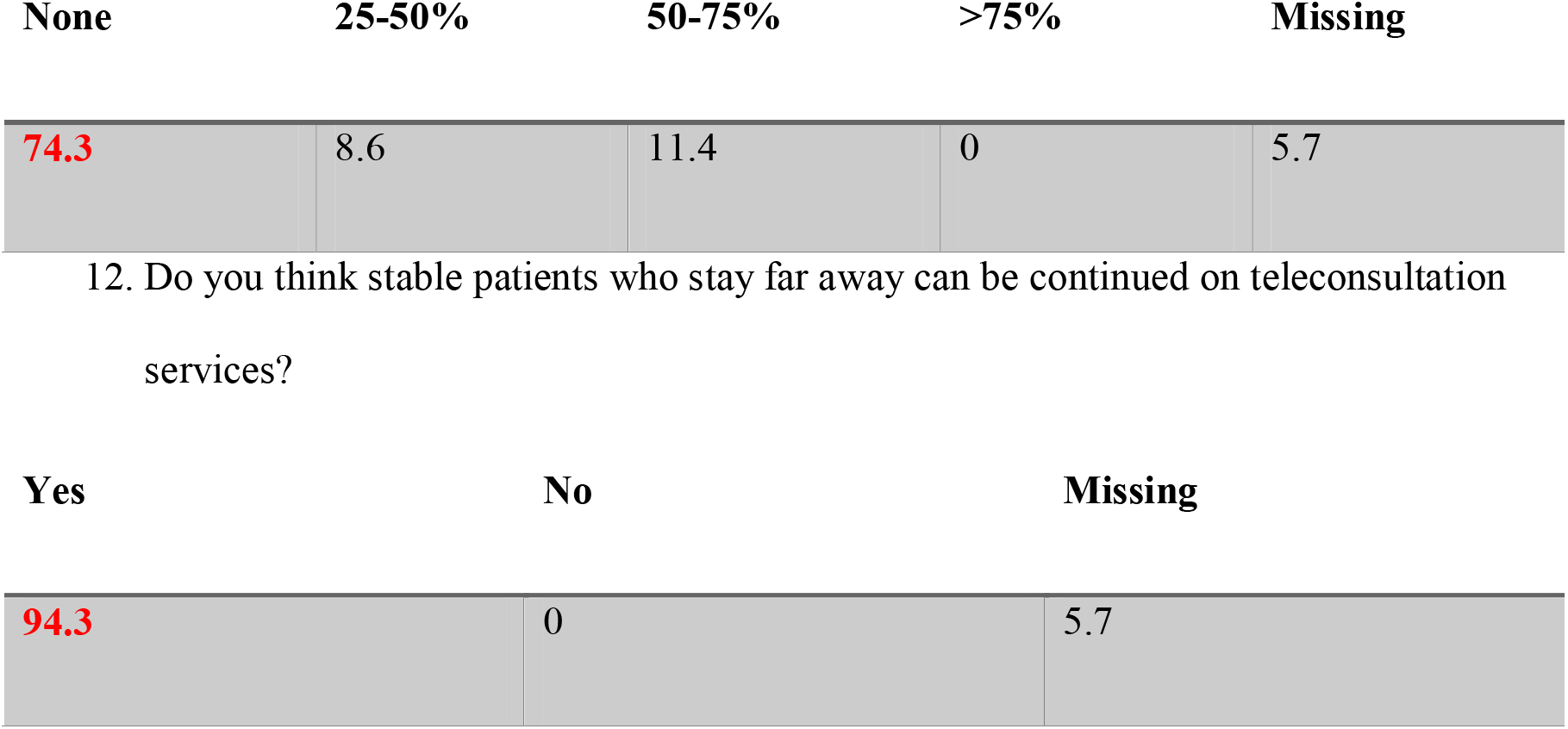
Questionnaire for Neurology faculty and trainees who conducted the tele-follow-up.

1. Was tele-follow-up useful (%)?
| Not at all | Fair | Good | Excellent | Unsure | Missing |
| --- | --- | --- | --- | --- | --- |
| 2.9 | 14.3 | 71.4 | 5.7 | 0 | 5.7 |

2. Were you able to fulfill the purpose of the conversation?
| Not at all | Fair | Good | Unsure | Missing |
| --- | --- | --- | --- | --- |
| 0 | 37.1 | 57.1 | 0 | 5.7 |

3. Did patients complain about difficulty in getting medications during lockdown?
| No | Rarely | Sometimes | Frequently | Always | Missing |
| --- | --- | --- | --- | --- | --- |
| 8.6 | 17.1 | 34.3 | 31.4 | 2.9 | 5.7 |

4. Were your patients continuing medications?
| <30% | 31-50% | 50-70% | 71-90% | >90% | Don't know | Missing |
| --- | --- | --- | --- | --- | --- | --- |
| 0 | 0 | 5.7 | 17.1 | 71.4 | 0 | 5.7 |

5. What do you think would be the satisfaction level of the patients with this teleconsultation?
| Unsatisfied | Little satisfied | Fairly satisfied | Completely satisfied | Missing |
| --- | --- | --- | --- | --- |
| 0 | 0 | 85.7 | 8.6 | 5.7 |

**To reassure To give medical advice To allay anxiety All Missing**
|  |  |  |  |  |
| --- | --- | --- | --- | --- |
| 5.7 | 11.4 | 0 | 77.1 | 5.7 |

**Not at all 50% 75% 100% Missing**
|  |  |  |  |  |
| --- | --- | --- | --- | --- |
| 0 | 11.4 | 68.6 | 14.3 | 5.7 |

**<25% 25-50% 50-75% >75% Missing**
|  |  |  |  |  |
| --- | --- | --- | --- | --- |
| 0 | 22.9 | 54.3 | 14.3 | 8.6 |

**<25% 25-50% 50-75% >75% Missing**
|  |  |  |  |  |
| --- | --- | --- | --- | --- |
| 2.9 | 11.4 | 62.9 | 14.3 | 5.7 |

**Not at all To some extent >50% of times Missing**
|  |  |  |  |
| --- | --- | --- | --- |
| 5.7 | 60.0 | 28.6 | 5.7 |

11. How many times did you use WhatsApp images/video of the patient/prescription?
| None | 25-50% | 50-75% | >75% | Missing |
| --- | --- | --- | --- | --- |
| 74.3 | 8.6 | 11.4 | 0 | 5.7 |

| Yes | No | Missing |
| --- | --- | --- |
| 94.3 | 0 | 5.7 |

## Discussion/Conclusion

Our study found that the tele-follow-up services were feasible and the level of satisfaction amongst Neurologists and trainees was good. The apprehension of tele-follow-up due to absence of conventional in-person meeting and examination was partly incorrect. While the lockdown left us with limited options to reach out to our patients, the availability of follow up online appointment list provided an opportunity to initiate and test this service. We could reach out to more than half of the scheduled appointments. Majority of the colleagues conducting the tele-follow-ups were satisfied with the utility and wanted to continue even after the lockdown for a select group of stable chronic neurological conditions where the patients were coming from distant locations.

The study showed how a crisis period proved to be a catalyst to initiate a neglected but useful service.^4^ It also provided information about the areas where we need to improve, including better documentation of phone numbers, streamlining of appointment system, addition of alternative methods of communication like emails, and even a prior notification of these calls to the patients, which was not done in this emergency situation.

Practice of telemedicine in specific areas of neurology like stroke^5–7^ and movement disorders^8–10^ have found reimbursement, legal considerations, and technological issues as the major barriers. The cost effectiveness of patients travels, stay and time spent on an in-person follow-up consultation needs to be weighed against both the physician and patient’s perspective. The most common response of our patients contacted via tele-follow-up was delight and intense gratitude. The communication transcended beyond the usual approach of medical history and examination as it also does in the clinic visits. Patients and their families in India look up to a physician more than a medical provider- a counselor, advisor, and even decision maker. The similar sentiment resonated in the tele-follow-up. Conversations ranged from concerns about their illness, enquiry about the pandemic, cautions to be taken and their concern and prayers for us.

The limitations were that the patient’s perceptions were not systematically recorded, as it could have contributed to the policy making for such facility in future. Another limitation could be a potential miscommunication but would only be known in subsequent visits. Making this a routine practice would require a mechanism of documentation of the conversation and prescription. It would additionally help in decongesting busy OPDs. Involvement of social workers and tele-rehabilitation will go a long way in improving services. A scheduled video interaction and email communication of prescription has barriers in a resource poor setting like India. Lack of literacy and smartphone use^11^ might be the possible challenges while putting it in routine use. We also did not use the video communication, and a prior intimation would have facilitated this in a sub-group of patients. This would have required more preparation and personnel.

## Conclusion

Tele-follow-up was feasible and satisfactory. While about half the patients could not be contacted, all faculty and trainees felt that tele-follow-up had the potential to be used for clinically stable patients living in distant locations.

## Data Availability

The anonymous data may be shared on request

## Statements

## Acknowledgement

None

## Statement of Ethics

Ethical clearance was taken from the Institute Ethics committee (IEC-301/17.04.2020).

## Author Contributions

This is to certify that **ALL** authors gave substantial contributions to the conception or design of the work; or the acquisition, analysis, or interpretation of data for the work; AND

Drafting the work or revising it critically for important intellectual content; AND

Final approval of the version to be published;

AND Agreement to be accountable for all aspects of the work in ensuring that questions related to the accuracy or integrity of any part of the work are appropriately investigated and resolved.

